# An optimal 1H-MRS technique at 7T: Proof-of-principle in Chronic Multiple Sclerosis and Neuromyelitis Optica Brain Lesions and Normal Appearing Brain Tissue

**DOI:** 10.1101/2020.07.17.20150730

**Authors:** George Tackley, Yazhuo Kong, Rachel Minne, Silvia Messina, Anderson Winkler, Ana Cavey, Rosie Everett, Gabriele C DeLuca, Andrew Weir, Matthew Craner, Irene Tracey, Jacqueline Palace, Charlotte J Stagg, Uzay Emir

**Author notes:** To whom correspondence should be addressed: Dr George Tackley, CUBRIC, Cardiff, Wales. Joint first authors (George Tackley, Yazhuo Kong). Joint last authors (Charlotte J Stagg, Uzay Emir).

## Abstract

Magnetic Resonance Spectroscopy (MRS) allows for the non-invasive quantification of neurochemicals and has the potential to differentiate between the pathologically distinct diseases, multiple sclerosis (MS) and AQP4Ab-positive neuromyelitis optica spectrum disorder (AQP4Ab-NMOSD). In this study we characterised the metabolite profiles of brain lesions in 11 MS and 4 AQP4Ab-NMOSD patients using an optimised MRS methodology at ultra-high field strength (7T) incorporating correction for T2 water relaxation differences between lesioned and normal tissue.

MS metabolite results were in keeping with the existing literature: total NAA was lower in lesions compared to normal appearing brain white matter (NAWM) with reciprocal findings for Inositol. An unexpected subtlety revealed by our technique was that total NAA differences were driven by NAA-glutamate (NAAG), a ubiquitous CNS molecule with functions quite distinct from NAA though commonly quantified together with NAA in MRS studies as total NAA. Surprisingly, AQP4Ab-NMOSD showed no significant differences for total NAA, NAA, NAAG or Inositol between lesion and NAWM sites, nor were there any differences between MS and AQP4Ab-NMOSD for *a priori* hypotheses. Post-hoc testing did however reveal greater total NAA in MS compared to AQP4Ab-NMOSD NAWM. Post-hoc testing also revealed a significant correlation between NAWM Ins:NAA and disability (as measured by EDSS) for disease groups combined, driven by the AP4Ab-NMOSD group.

Utilising an optimised MRS methodology, our study highlights some under-explored subtleties in MRS profiles, such as the absence of Inositol concentration differences in AQP4Ab-NMOSD brain lesions versus NAWM and the important influence of NAAG differences between lesions and normal appearing white matter in MS.

## 1. Introduction

Magnetic Resonance Spectroscopy (MRS) allows for the non-invasive quantification of neurochemicals, and therefore has the potential to differentiate between pathologically distinct diseases. This is perhaps especially important in circumstances where clinical syndromes may overlap but treatment strategies are distinct. In particular, there is interest in utilising MRS to differentiate the primary astrocytopathy Aquaporin-4 Antibody positive Neuromyelitis Optica Spectrum Disorders (AQP4Ab-NMOSD) (Fujihara, 2011) from the clinically similar but pathologically distinct disorder, Multiple Sclerosis (MS), which is believed to be a chronic inflammatory demyelinating disorder with secondary neurodegeneration (Lucchinetti et al., 2014; Wingerchuk et al., 2015). In addition, where MRS metrics prove sensitive to the underlying pathology in a disease state, they can inform our understanding of that pathology, and potentially be developed as biomarkers for future pharmacological studies.

MRS studies in MS consistently report a core pattern of findings. Compared to healthy control brain tissue, lesions of all ages show reduced total N-acetyl aspartate (tNAA) likely reflecting decreased neuronal mitochondrial activity, and raised Inositol-containing compounds (Ins), commonly taken to reflect glial activity (Brex et al., 2000; Ciccarelli et al., 2013; Miller et al., 2003). A similar pattern of changes is found in MS normal appearing white matter (NAWM) versus healthy controls, though to a smaller degree (De Stefano et al., 2007; Fernando et al., 2004; Helms et al., 2000; Miller et al., 2003). In addition to changes in specific metabolites, the relative concentrations of some MRS metabolites have also been correlated with clinical metrics. The relative concentration of Inositol and NAA (without NAAG) in NAWM (Ins:NAA), which provides an insight into the relative activity of glia and neurons within a given region, has been shown to predict disease progression in MS. It has been hypothesised to be a sensitive index that encapsulates both the damaging immune-mediated gliosis (increase in Inositol) and the disabling neurodegenerative axonal loss (reduction in NAA) that are known features of MS pathology, and it has been shown to predict disease progression (Llufriu et al., 2014; Miller DH, 2014).

In patients with AQP4Ab-NMOSD, there is little or no evidence for a difference in either tNAA (i.e. NAA + NAAG) or Inositol in normal appearing white matter compared to controls (Aboul-Enein et al., 2010; Bichuetti et al., 2008; Kremer et al., 2015), raising the possibility that Inositol and tNAA may be sensitive discriminators between AQP4Ab-NMOSD and MS. However, no studies to date have studied MRS-quantified neurochemicals within AQP4Ab-NMOSD lesions in the brain. A single study has examined MRS-derived neurochemical profiles in AQP4Ab-NMOSD lesions within the spinal cord and demonstrated significantly lower Inositol in AQP4Ab-NMOSD lesions compared with both MS cord lesions and healthy controls (Ciccarelli et al., 2013), consistent with previously observed astrocyte damage and loss in AQP4Ab-NMOSD lesions. The authors also demonstrated, in line with existing literature, a substantial decrease in tNAA in MS lesions compared with healthy controls, but no significant difference in tNAA between AQP4Ab-NMOSD lesions and either MS lesions or controls.

Here, we wished to see whether MRS could distinguish between AQP4Ab-NMOSD and MS lesions in the brain. However, there are a number of technical difficulties in performing MRS in the context of neurological disease that are important to overcome in order to accurately address this question. In particular, lesioned tissue has a longer T2 than non-lesioned tissue, due to its increased water content (Zimmerman et al., 1986), which, if not compensated for (e.g. by using a short TE and T2* measurement), will directly influence metabolite quantification. In addition, it is important to fully separate neurochemicals with highly similar spectral resonances but disparate physiological functions, for example NAA and the closely chemically related, but functionally distinct, NAA-glutamate (NAAG), often combined in the MRS metric tNAA. This is particularly important in the context of neurodegeneration, as NAA reflects neuronal mitochondrial function, whereas NAAG acts as a neuromodulator (Baslow, 2000; Birken and Oldendorf, 1989; Neale et al., 2000). It is therefore difficult to clearly interpret changes in the tNAA metric in terms of the underlying pathology.

We therefore exploited recent advances in MRS methodology to study pathological differences between AQP4Ab-NMOSD and MS lesions. We used an ultra-high field (7T) scanner, to allow separation between NAA and NAAG, something not easily achievable at 3T, and modelled multiple water T2-relaxation times to compensate for lesion-related effects on metabolite quantification (Helms, 2001), approaches that have been little used in this context before now.

We wished to test the hypotheses that (1) Inositol would be higher in MS brain lesions than AQP4Ab-NMOSD brain lesions, reflecting the likely increased astrocytic damage in AQP4Ab-NMOSD compared to the reciprocal astrogliosis found in MS lesions, and (2) that NAA (tNAA, NAA & NAAG) would be higher in the normal appearing white matter in AQP4Ab-NMOSD patients compared with MS patients, in line with the relative lack of extra-lesional neurodegeneration in AQP4Ab-NMOSD (Matthews et al., 2015). Within diseases we also hypothesised that (3) NAA (tNAA, NAA & NAAG) would be greater in NAWM than lesion sites in line with expected neuronal loss in lesions, and that (4) inositol would be differentially *greater* in MS brain lesion versus NAWM sites and lower in AQP4Ab-NMOSD lesion versus NAWM sites reflecting the contrasting gliotic and astrocytopathic nature of lesions in these two conditions.

## 2. Methods

### 2.1 Subjects

Eleven patients with clinically diagnosed relapsing-remitting multiple sclerosis (RRMS) and four patients with AQP4Ab-NMOSD gave their written informed consent to participate in the study, under local ethics board approval (Oxfordshire REC A 10/H0604/99; Berkshire REC 13/SC/0238). In addition to MR scanning, patients underwent a short clinical consultation and neurological examination including EDSS scoring. Current medications were recorded. Minimum lesion age was calculated as the time from the oldest clinical brain MRI to contain the targeted lesion.

### 2.2 MR Acquisition

MR was performed on a 7T Siemens MAGNETOM system (Siemens, Erlanden, Germany) equipped with a Nova Medical 32 channel receive array head coil. Two MRS volumes-of-interest (VOIs) were acquired per subject: one targeting a chronic, T2-hyperintense, T1-hypointense white matter lesion (>3 months old, confirmed on historical clinical MRIs) and another centred on a contralateral area of normal appearing white matter (NAWM; a majority white-matter voxel, avoiding as much grey matter and cerebrospinal fluid as possible), positioned as close as possible to contralateral reflection-symmetrical with the lesion voxel. The spectroscopy voxel volume was 15 × 15 × 15mm.

Spectroscopy voxels were manually positioned by reference to a 1mm isotropic T2-weighted fluid-attenuated inversion recovery (FLAIR) image (1mm isotropic, TR = 5s, TE = 272 ms, TI = 1.8 sec) (See Supplemental Figure 1 for individual voxel locations). First- and second-order shims were first adjusted by gradient-echo shimming (Shah et al., 2009). The second step involved only fine adjustment of first order shims using FASTMAP (Gruetter and Tkáč, 2000). Spectra were acquired using a Stimulated Echo Acquisition Mode (STEAM) pulse sequence (TE=11ms, TR=5s, number of transients=64) with variable power radiofrequency pulses with optimized relaxation delay (VAPOR), water suppression and outer volume saturation (Emir et al., 2012). Unsuppressed water spectra acquired from the same voxel were used to remove residual eddy current effects and to reconstruct the phased array spectra.

Finally, fully relaxed unsuppressed water signals were acquired at TEs ranging from 11 to 4000ms (TR=15s) to estimate the cerebrospinal fluid (CSF) contribution to each VOI (see below). A whole-brain T1-MPRAGE (1mm isotropic, TR = 2.2s, TE = 282ms) was also acquired to evaluate T1 hypo-intensity in lesions.

### 2.3 MRS Analysis

Absolute metabolite concentrations were obtained relative to an unsuppressed water spectrum acquired from the same VOI. The transverse relaxation times (T2) of tissue water and percent CSF contribution to the VOI were obtained by fitting the integrals of the unsuppressed water spectra acquired in each VOI at different TE values with a biexponential fit (Piechnik et al., 2009), with the T2 of CSF fixed at 640ms and three free parameters: T2 of tissue water, amplitude of tissue water, and amplitude of CSF water.

LCModel (Provencher, 2001) was used for spectral analysis and quantification. Single-subject metabolites were only retained if the Cramér-Rao lower bounds (CRLBs) estimated error of metabolite quantification was less than 20%, and average metabolites were only reported where at least 50% of single-subject measurements met this criterion.

### 2.4 Statistics

Cohort characteristics of age, disease duration, EDSS and minimum age of lesions were compared using unpaired t-tests; sex and ethnicity were compared using chi-square with Yate’s correction. Paired t-tests were used to compare mean T2-water estimates and measures of spectral quality. All subsequent group-level statistics were computed using permutation testing in FSL’s PALM with 10,000 permutations (Alberton et al., 2020; Jenkinson et al., 2012; Winkler et al., 2016). Unpaired two-sample t-tests were used to compare lesions and areas of NAWM across groups (hypotheses 1 and 2). One-sample t-tests were performed to test individual lesion and NAWM metabolite concentration differences within disease groups (hypotheses 2 and 3). We ran statistical tests for metabolite comparisons in three batches (the fewest number that data structure and test methodology would allow) to allow for family wise error rate correction. The three batches were: unpaired t-tests for MS and AQP4Ab-NMOSD comparisons, one-sample t-test for AQP4Ab-NMOSD lesion versus NAWM differences and one-sample t-tests for MS lesion versus NAWM differences. In order to test our directionally specific hypotheses, all *a priori* group-level t-tests were one-tailed. Post-hoc two-tailed t-tests, as detailed in the results, were also performed in FSL PALM.

Tests of correlation were limited to common associations described in the literature, namely disability’s association with NAA:tCr (normal appearing brain tissue & lesions) (Khan et al., 2016; Mainero et al., 2001), Ins:NAA (normal appearing white and grey matter) (Llufriu et al., 2014; Miller DH, 2014) and GABA (normal appearing grey matter) (Cawley et al., 2015), and disease duration’s association with Ins, NAA, Cr, tCr and tCho (normal appearing brain tissue) (Kirov et al., 2013). Correlations were evaluated visually and with R-square (R^2^), however due to low numbers of participants and the relatively large number of tests, p-values were not calculated except for post-hoc testing of Pearson’s correlation coefficient in the case of mI:NAA vs EDSS (See Supplemental figure 2). Pearson’s correlations performed in R (R Core Team, 2017; Wickham, 2016).

## 3. Results

Patient characteristics are listed in Table 1. Three of the four AQP4Ab-NMOSD participants were Afro-Caribbean and one was Asian, in line with the non-Caucasian predominance in this disease. One MS participant was Afro-Caribbean, the remainder were Caucasian. All lesions studied were hyper-intense on the FLAIR image and hypo-intense on the T1 weighted image. The minimum age of the brain lesions ranged from 132 days to 9 years.

**Table 1.** Demographics and clinical features

|  | MS | NMO | All |
| --- | --- | --- | --- |
| Total n | 11 | 4 | 15 |
| Age, yrs, median (min-max) | 43 (28-60) | 38 (24-40) | 40 (24-60) |
| Female, n (%) | 7 (63.6) | 3 (75) | 10 (66.7) |
| Race |  |  |  |
| Afro-caribbean (%) | 1 (9.1)** | 3 (75)** | 4 (26.7) |
| Caucasian (%) | 10 (90.9)** | 0 (0)** | 10 (66.7) |
| Asian (%) | 0 (0)** | 1 (25)** | 1 (6.7) |
| Disease duration, yrs, mean (sd) | 6.8 (5.5) | 8.9 (3.2) | 7.3 (5.1) |
| EDSS, median (min-max) | 2 (0-6) | 3.75 (0-6) | 2.5 (0-6) |
| Minimum age of lesion, months, mean (sd) | 36 (33.3)* | 86.6 (27.1)* | 49.5 (38.9) |
| On treatment, n (%) | 8 (72.7) | 4 (100) | 12 (80) |

\* p<0.05 for MS vs NMO t-test comparison
\*\* p<0.05 for MS vs NMO chi-square comparison

### 3.1 MRS quality metrics

We first wanted to ensure that there were no systematic differences in the quality of the LCModel fit between the NAWM and lesion VOIs. Reported CRLB estimates were generally low in all cases. However, in our MS sample Ala, Asc, Asp, Lac, PCho and Scyllo did not meet our CRLB criterion and were therefore excluded from further analysis. Likewise, for MS, 5 lesion and 5 NAWM measures were excluded from GABA analysis, 1 lesion and 4 NAWM measures from Gln, 1 lesion and 1 NAWM measure from Glc+Tau, 2 lesion and 1 NAWM measure from PE, and 1 NAWM measure from Tau. For AQP4Ab-NMOSD data, Ala, Asp, Lac, PCho and Scyllo did not meet the CRLB requirements and were excluded. Likewise, 1 lesion measure was excluded from GABA analysis and 2 lesion and 2 NAWM measure from Asc. For the remaining metabolites we then wished to ensure that there was no significant difference in the quality of fit between NAWM and lesion groups. Statistical analyses demonstrated no differences, when tests were corrected for multiple comparisons (paired t-test p-values, α = 0.05, Bonferroni threshold = 0.05/19 = 0.0026: 0.0025 < p < 0.05: AQP4Ab-NMOSD Gln p=0.011, AQP4Ab-NMOSD Asc 0.015, MS Glu 0.049; p > 0.05 for all other metabolites; see Table 3).

**Table 3A.** Multiple sclerosis metabolite profile (n=11)

| *Metabolite* | Concentration, mmol/L |  |  |  | CRLB |  |
| --- | --- | --- | --- | --- | --- | --- |
|  | *Lesion,*<br>*mean (sd)* | *n* | *NAWM,*<br>*mean (sd)* | *n* | *Lesion,*<br>*mean (sd)* | *NAWM,*<br>*mean (sd)* |
| Cr | 3.67 (0.3) | 11 | 3.69 (0.3) | 11 | 8.73 (1.5) | 8.55 (1.2) |
| PCr | 4.19 (0.5) | 11 | 4.11 (0.5) | 11 | 8 (1.3) | 8.09 (0.9) |
| tCr | 7.85 (0.7) | 11 | 7.79 (0.7) | 11 | 2.27 (0.4) | 2.09 (0.3) |
| GABA | 1.27 (0.3) | 6 | 1.32 (0.1) | 6 | 16.5 (3.1) | 16 (1.2) |
| Glc+Tau | 1.8 (0.5) | 10 | 1.61 (0.4) | 10 | 12.5 (2.6) | 12.7 (2) |
| Gln | 2.1 (0.5) | 10 | 2.06 (0.5) | 7 | 12.4 (2.4) | 13.29 (3.4) |
| Glu | 6 (0.8) | 11 | 6.61 (1.1) | 11 | 4.27 (1) | 3.64 (0.6) |
| Glx | 8.04 (1.1) | 11 | 8.34 (1.5) | 11 | 4.45 (0.9) | 4.36 (0.8) |
| GPC | 1.68 (0.3) | 11 | 1.63 (0.4) | 11 | 6 (1.7) | 6.36 (2.1) |
| tCho | 1.91 (0.4) | 11 | 1.78 (0.3) | 11 | 4.09 (0.8) | 4.18 (0.8) |
| GSH | 1.22 (0.2) | 11 | 1.18 (0.2) | 11 | 10.55 (1.6) | 10.91 (1.8) |
| Ins | 8.1 (1.4) | 11 | 7.28 (1.2) | 11 | 2.73 (0.6) | 3.09 (0.3) |
| NAA | 8.94 (1.1) | 11 | 9.69 (0.9) | 11 | 2.55 (0.5) | 2.36 (0.5) |
| NAAG | 2.09 (0.6) | 11 | 2.55 (0.9) | 11 | 8 (2.5) | 7 (2.9) |
| tNAA | 11.03 (1.1) | 11 | 12.24 (0.9) | 11 | 1.91 (0.3) | 1.91 (0.3) |
| PE | 1.89 (0.4) | 9 | 2.18 (0.4) | 10 | 15.89 (2.7) | 14.2 (2.1) |
| Tau | 1.66 (0.4) | 11 | 1.6 (0.4) | 10 | 12.82 (2.6) | 12.6 (2) |

NAWM, normal appearing white matter; CRLB, Cramér–Rao lower bound

### 3.2 T2 differences between lesions and normal-appearing tissue are relevant for metabolite quantification

There was no difference between the LCModel estimated line-widths (Full Width Half Maximum, FWHM) and Signal-to-Noise ratios (S/N) of the spectra from the NAWM and lesioned tissue (table 2; S/N: 25.7±5.6 vs 28.3±4, p=0.10; FWHM: 0.03±0 vs 0.03±0, p=0.96). However, estimated T2-water relaxation time was higher in both MS and AQP4Ab-NMOSD lesion voxels than for NAWM voxels, reflecting the higher free-water content in this tissue (43.7±2.8 vs 39.9±2.6, p=0.0001). We therefore went on to quantify neurochemicals in MS and AQP4Ab-NMOSD lesions and NAWM spectra using T2-corrected spectra.

**Table 2.** Estimated T2-water, LCmodel estimated FWHM and SNR

|  | **Lesion** | **Control** | **p** |
| --- | --- | --- | --- |
| T2 water (ms) | 43.7 (2.8) | 39.9 (2.6) | 0.0001 |
| FWHM (ppm) | 0.03 (0) | 0.03 (0) | 0.958 |
| S/N | 25.7 (5.6) | 28.3 (4) | 0.097 |

FWHM, full width half maximum
S/N, signal to noise ratio

### 3.3 Differences in neurochemical profiles between NAWM and lesions in MS

Next, we investigated neurochemical differences between lesions and NAWM. All *a priori* group-level t-tests were unidirectional (one-tailed) in accordance with our hypotheses. In the MS group absolute tNAA was lower in lesions compared to NAWM ([NAWM tNAA] – [lesion tNAA]: 1.21 ±1.31 (mean ± SD); one-sample t(10)=2.91, p=0.020). Given that, as discussed above, tNAA is comprised of two metabolites with very different physiological roles, we therefore wanted to investigate whether this difference in tNAA was driven by a decrease in NAAG, or NAA, or both. NAAG was lower in lesions than in NAWM ([NAWM NAAG] – [lesion NAAG]: 0.46 ±0.57; one-sample t(10)=2.52, p=0.048), but only a trend towards lower values in lesions was found for NAA ([NAWM NAA] – [lesion NAA]: 0.75 ±1.13; one-sample t(10)=2.10, p=0.076) (Figures 2 & 3 and Table 3A). Again, Inositol was greater in MS lesions than in NAWM ([lesions Ins] – [NAWM Ins]: 0.82 ±0.99; t(10)=2.62; p=0.036; Figure 3 & Table 3A).

**Figure 1.**
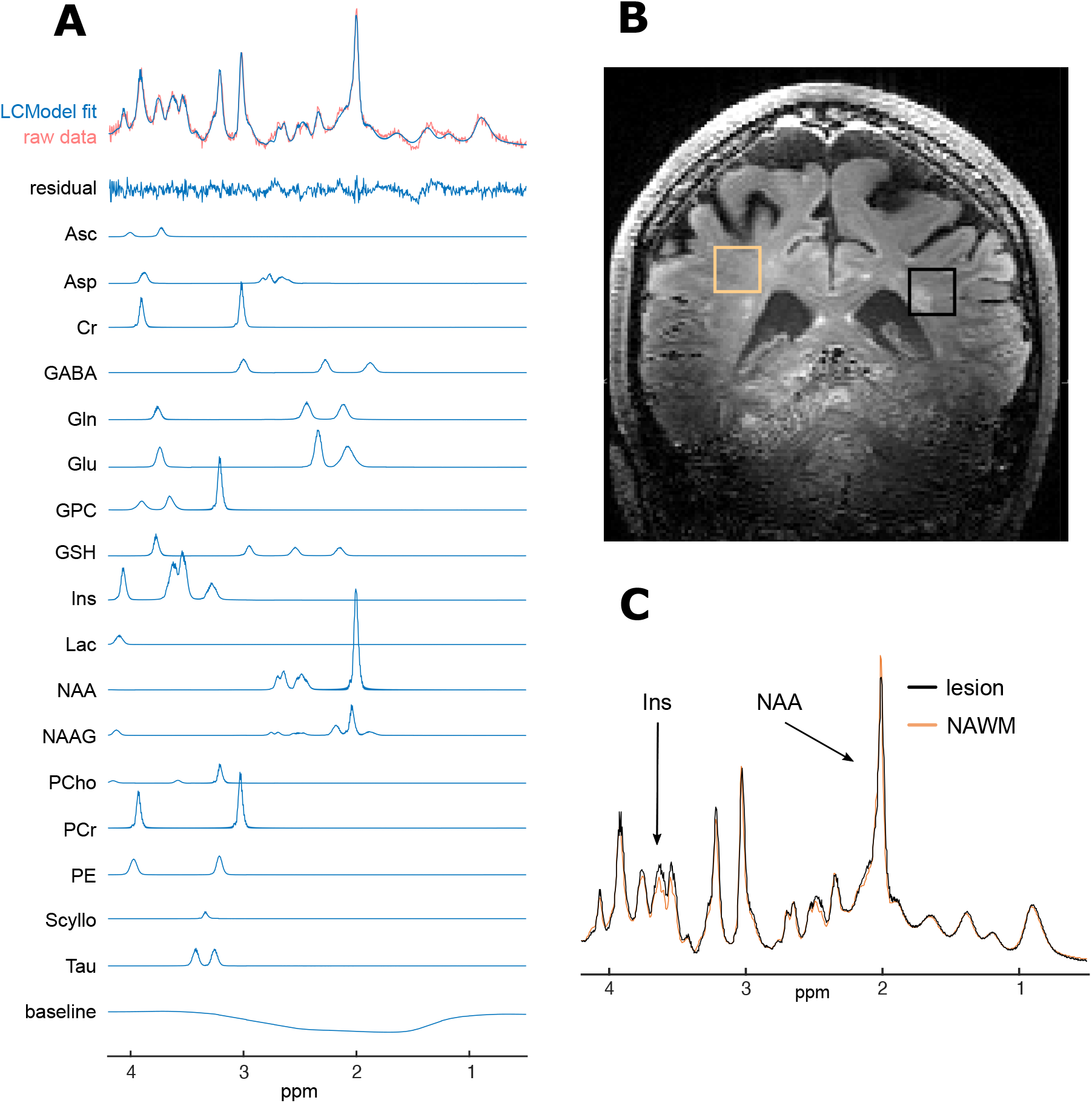
Example spectra and voxel placement from single subject. A. Raw spectrum data, LCModel fit (for average spectra and individual metabolites), residual-error and baseline for example participant (no. 20); B. Example voxel placement (lesion outlined in black and NAWM region outlined in orange) for same subject; C. Comparison of fitted (and baseline subtracted) lesion and NAWM spectra showing differences in mI and NAA peaks, again for same subject. ppm, parts per million; Ins, *myo*-inositol; NAA, N-aspartylaspartate.

**Figure 2.**
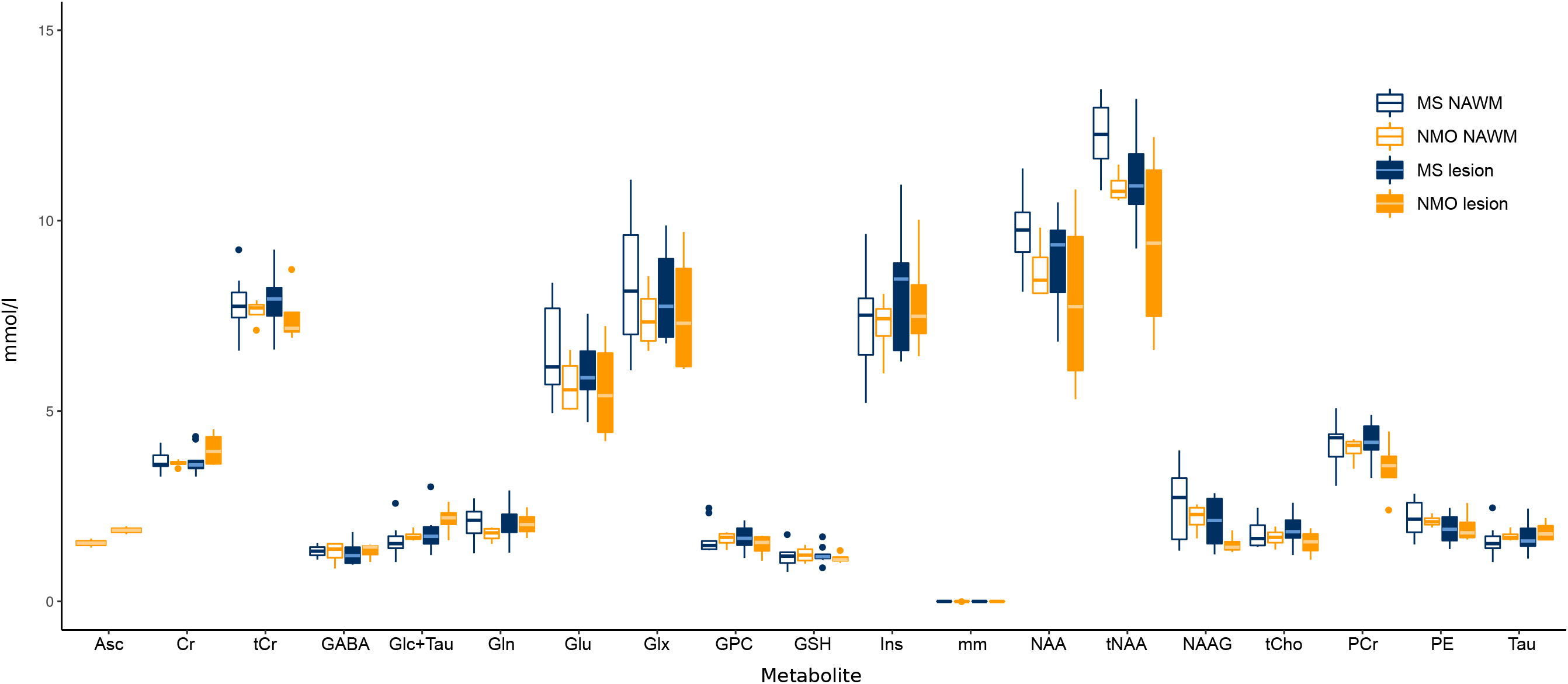
Metabolite profiles for multiple sclerosis and AQP4Ab positive neuromyelitis optica. Box and whisker plots of metabolites for MS and AQP4Ab-NMOSD (NMO; min, lower-quartile, median, upper-quartile, max; outliers are >1.5 × interquartile range beyond lower or upper quartiles). MS, multiple sclerosis; NMO, AQP4-antibody positive neuromyelitis optica; NAWM, normal appearing white matter

**Figure 3.**
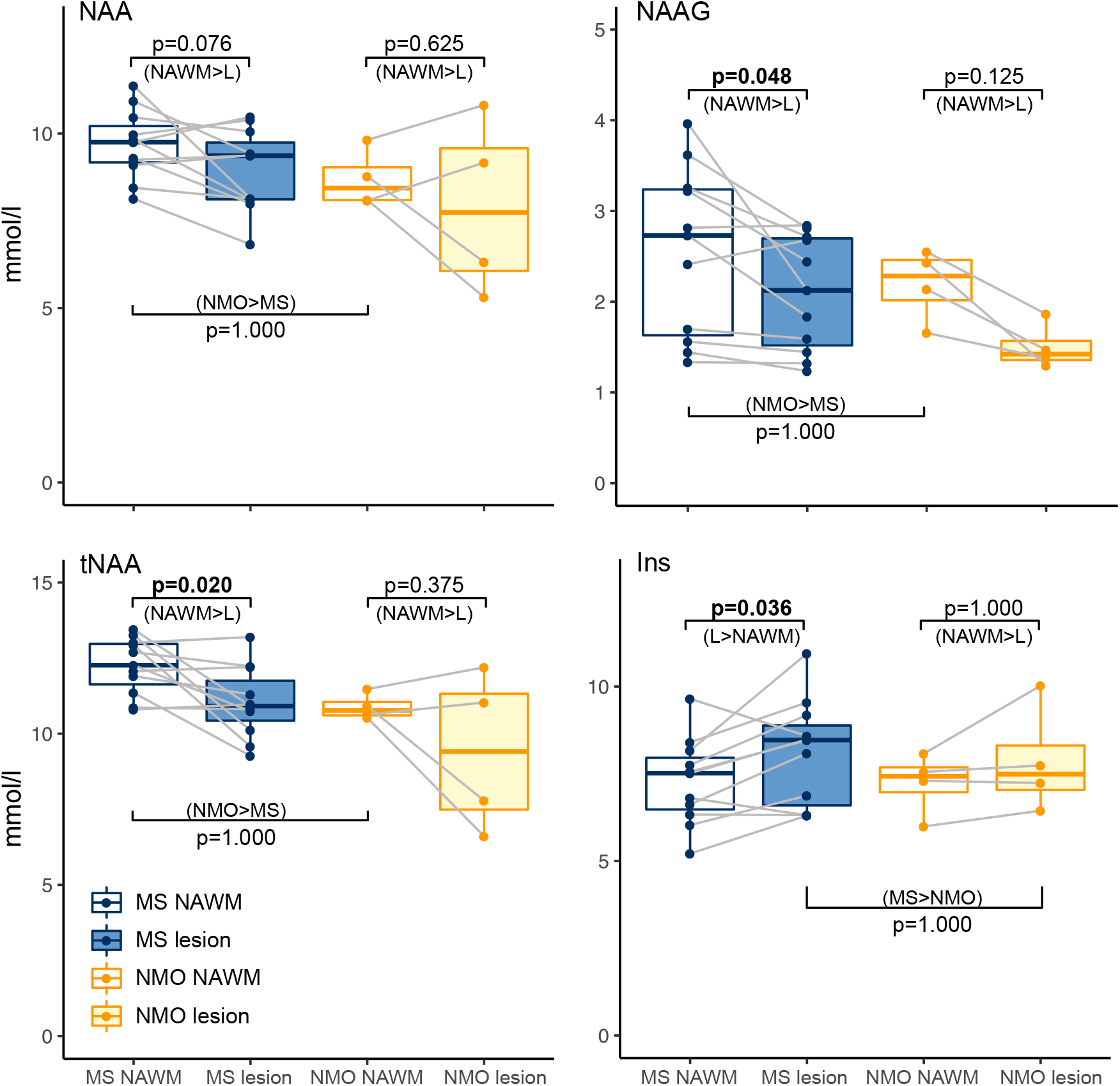
Individual metabolite comparison boxplots. Boxplot representations of metabolite comparisons based on *a priori* hypotheses. Hypotheses were unidirectional hence p-values denote significance of one-tailed t-tests. Direction of test denoted in brackets beneath p-value. MS, multiple sclerosis; NMO, neuromyelitis optica; NAWM, normal appearing white matter; L, lesion. (Figure produced with R ggplot2 package; Wickham, 2016).

### 3.4 No difference in neurochemical profiles between NAWM and lesions in AQP4Ab-NMOSD

Again with one-tailed t-tests of NAWM and lesion metabolite concentration differences, within the AQP4Ab-NMOSD patients, we found no difference in tNAA (note that this is n=4; [NAWM tNAA]- [lesion tNAA]: 1.48 ±2.06 (mean ± sd); one-sample t(3)=1.24, p=0.375). When the two metabolites that contribute to tNAA were investigated separately, neither NAA nor NAAG differed between NAWM and lesions, despite NAAG being visibly greater in NAWM ([NAWM NAA]–[lesion NAA]: 0.79 ±1.84; one-sample t(3)=0.74, p=0.625; [NAWM NAAG]-[lesion NAAG]: 0.69 ±0.3; one-sample t(3)=3.96, p=0.125) (See Figure 3). Inositol did not differ between NAWM and lesioned tissue ([NAWM Ins] – [lesion Ins]: −0.63 ±0.78; one-sample t(3)=-1.40, p=1.000) (see Figure 3 & Table 3B). There were thus no differences in either tNAA, NAA, NAAG or Inositol between the lesion and NAWM groups in AQP4Ab-NMOSD.

**Table 3B.** AQP4-Ab positive neuromyelitis optica spectrum disorder metabolite profile (n=4)

| *Metabolite* | Concentration, mmol/L |  |  |  | CRLB |  |
| --- | --- | --- | --- | --- | --- | --- |
|  | *Lesion,*<br>*mean (sd)* | *n* | *NAWM,*<br>*mean (sd)* | *n* | *Lesion,*<br>*mean (sd)* | *NAWM,*<br>*mean (sd)* |
| Cr | 4 (0.4) | 4 | 3.63 (0.1) | 4 | 7.75 (0.8) | 9 (1.4) |
| PCr | 3.5 (0.7) | 4 | 3.98 (0.3) | 4 | 9.5 (2.7) | 8.75 (1.3) |
| tCr | 7.5 (0.7) | 4 | 7.61 (0.3) | 4 | 2 (0) | 2.25 (0.4) |
| GABA | 1.32 (0.2) | 3 | 1.28 (0.3) | 4 | 15.67 (2.6) | 16.75 (2.6) |
| Glc+Tau | 2.15 (0.4) | 4 | 1.72 (0.1) | 4 | 11 (1.6) | 11 (2.9) |
| Gln | 2.04 (0.3) | 4 | 1.76 (0.2) | 4 | 11.5 (1.1) | 14.25 (1.1) |
| Glu | 5.56 (1.2) | 4 | 5.69 (0.7) | 4 | 4 (0.7) | 4.5 (0.9) |
| Glx | 7.6 (1.5) | 4 | 7.45 (0.8) | 4 | 4 (0.7) | 4.5 (0.9) |
| GPC | 1.47 (0.3) | 4 | 1.63 (0.2) | 4 | 5.5 (2.1) | 5.75 (1.8) |
| tCho | 1.54 (0.3) | 4 | 1.67 (0.2) | 4 | 4.5 (1.1) | 4.5 (0.5) |
| GSH | 1.14 (0.1) | 4 | 1.23 (0.2) | 4 | 10.5 (0.9) | 10.25 (1.6) |
| Ins | 7.86 (1.3) | 4 | 7.23 (0.8) | 4 | 2.75 (0.4) | 3 (0) |
| NAA | 7.9 (2.2) | 4 | 8.69 (0.7) | 4 | 2.5 (0.5) | 2.5 (0.5) |
| NAAG | 1.5 (0.2) | 4 | 2.19 (0.3) | 4 | 9.25 (2.9) | 7.5 (2.7) |
| tNAA | 9.4 (2.3) | 4 | 10.89 (0.4) | 4 | 2 (0) | 1.75 (0.4) |
| PE | 1.95 (0.4) | 4 | 2.11 (0.1) | 4 | 14.25 (1.5) | 13.75 (2.7) |
| Tau | 1.84 (0.2) | 4 | 1.72 (0.1) | 4 | 11 (1.6) | 11 (2.9) |
| Asc | 1.87 (0.1) | 2 | 1.53 (0.1) | 2 | 12.5 (0.5) | 16.5 (0.5) |

NAWM, normal appearing white matter; CRLB, Cramér–Rao lower bound

### 3.5 Greater tNAA in NAWM in MS compared with AQP4Ab-NMOSD

We then wanted to compare neurochemical concentrations between MS and AQP4Ab-NMOSD. Unpaired one-tailed two-sample t-tests were used. NAWM concentrations of NAA, NAAG and tNAA were not lower in MS NAWM (AQP4Ab-NMOSD>MS NAWM NAA: 8.69 ±0.7 > 9.69 ±0.9; two-sample t(13)=-1.7912; AQP4Ab-NMOSD>MS NAWM NAAG: 2.19 ±0.3 > 2.55 ±0.9; two-sample t(13)=-0.7316; AQP4Ab-NMOSD>MS NAWM tNAA: 10.89 ±0.4 > 12.24 ±0.9; two-sample t(13)=-2.7458; all p-values >1.000). However, on visual inspection of the data, tNAA appeared to be greater in MS compared to AQP4Ab-NMOSD NAWM. A post-hoc two-tailed unpaired t-test (uncorrected for multiple comparisons) was significant (MS>AQP4Ab-NMOSD tNAA: 12.24 ±0.9 > 10.89 ±0.4; t(13)=2.75, p=0.0183).

Inositol was not greater in MS versus AQP4AB-NMOSD lesions (MS>AQP4Ab-NMOSD Ins: 8.1 ±1.4 > 7.86 ±1.3; two-sample t(13)=0.268, p=1.000) (Figure 3).

For completeness, we tested for a significant interaction of site (NAWM / lesion) by group (MS / AQP4Ab-NMOSD) for NAA, NAAG, tNAA and Ins (including our *a priori* expectations regarding the direction of difference) and this was non-significant for all comparisons (all p>0.9).

### 3.6 Ins:NAA correlates with clinical score in AQP4Ab-NMOSD

Finally, we wished to investigate whether there were any relationships between our metabolites of interest and disease scores (See Supplemental figure 2). Of relationships previously reported in the literature, we demonstrated a striking linear correlation between NAWM Ins:NAA and EDSS for both MS and AQP4Ab-NMOSD (MS: R^2^=0.22, AQP4Ab-NMOSD: R^2^=0.91, respectively; R^2^=0.30 combined). Post-hoc tests, uncorrected for multiple comparisons, showed that this was significant for the pooled data (AQP4Ab-NMOSD + MS NAWM Ins:NAA ∝ EDSS: r(13)=0.55, p=0.033) and for AQP4Ab-NMOSD alone (AQP4Ab-NMOSD NAWM Ins:NAA ∝ EDSS; r(2)=0.95; p=0.048), but not for MS alone (MS NAWM Ins:NAA ∝ EDSS: r(9)=0.47, p=0.141).

We additionally explored the relationship between Ins:NAA, disease group (MS / AQP4Ab-NMOSD) and MRS site (lesion / NAWM) with post-hoc two-tailed t-tests (uncorrected for multiple comparisons). We found that lesion Ins:NAA was significantly greater than NAWM Ins:NAA in MS (MS lesion Ins:NAA vs control Ins:NAA: 0.76 ±0.12 vs 0.91 ±0.10; one-sample t(10)=5.42) but for all other comparisons was non-significant.

## 4. Discussion

This study was performed in order to investigate the patterns of neurochemical changes in lesioned tissue and in normal appearing white matter in multiple sclerosis (MS) and aquaporin-4 antibody positive neuromyelitis optica spectrum disorder (AQP4Ab-NMOSD). To do this, we acquired Magnetic Resonance Spectroscopy (MRS) data using an optimised sequence for seven tesla (7T) MRI. We demonstrated significant differences in water T2 relaxation times between lesions and NAWM, which we corrected for in subsequent analyses.

Using this corrected data, as expected, Total NAA (tNAA) was lower in MS lesions compared to MS NAWM and Inositol was greater in MS lesions compared to MS NAWM, in line with historical MRS studies that attribute these features respectively to the axonal loss and gliosis seen in MS lesions (Arnold et al., 1992; Davie et al., 1994). We demonstrated no differences between lesion and NAWM in AQP4Ab-NMOSD patients, and no differences in either NAA or Inositol between MS and AQP4Ab-NMOSD lesions. We did show, however, a relationship between NAA:Ins, commonly thought to be a marker of neuronal loss and gliosis, and clinical disability score in both groups (Llufriu et al., 2014; Miller DH, 2014).

### 4.1 Optimised MRS allowed us to address important confounds

A number of parameters were optimised to acquire our 7T MRS spectra. The STEAM sequence was chosen despite the loss of half of the available signal to minimize relaxation effects at a short echo time. Transverse relaxation differences between lesion sites and NAWM at longer TEs have the potential to confound the quantification of metabolite concentrations.

The achieved spectral quality (high spectral resolution, SNR, efficient water suppression, and a distortionless baseline) allowed reliable quantification of 17-18 metabolites in periventricular white matter (VOI = 15 × 15 × 15 ml) using LCModel analysis. T2-water relaxation times were significantly higher in lesioned tissue as confirmed in previous studies (Laule et al., 2007b, 2007a), and quantifies what one would expect given their features on T2-weighted images where lesions are identified clinically by their bright (hyperintense) appearance.

We avoided over-reliance on metabolite ratios (e.g. tNAA:tCr) and corrected for multiple T2-relaxation components. Both are of particular importance within neuroinflammatory conditions, the former because a parallel loss of tCr in damaged tissue may conceal tNAA loss when expressed as tNAA:tCr (Davies et al., 1995) and the latter because brain lesions often have a higher percentage of water compared to surrounding tissue. Accurate absolute quantification of metabolites, powerful in their own right, also allow for meaningful interpretation of metabolite ratios where applied.

### 4.2 Lower NAA-G in MS lesions than in NAWM

A decrease in NAA in MS lesions has long been described (Arnold et al., 1992; Davie et al., 1994). However, a number of questions have remained to be conclusively answered about this finding, which we are able address here. Firstly, as discussed above, accounting for T2 relaxation times allows us to be sure that this is a true reflection of the underlying pathology and not an artefact of the acquisition protocols.

Secondly, our optimised MRS allowed us to accurately separate tNAA into its constituent parts: NAA and NAA-G. Although these two neurochemicals have similar molecular structures, meaning that they are hard to distinguish using MRS, they have distinct functional roles. The role of NAA is not entirely clear, but is a reflection of neuronal mitochondrial function, and has been hypothesised to have a role in myelination (Birken and Oldendorf, 1989; Moffett et al., 2007). NAAG, the most abundant peptide in the central nervous system, is found in both neurones and glia, acts as both a neurotransmitter and a glutamate reservoir, and is higher in white matter compared to grey matter (Chiew et al., 2018; Neale et al., 2000). NAAG in MS NAWM may reflect glial cell number and may also have a neuroprotective role via its ability to activate the metabotropic glutamate receptor, mGluR. As discussed above, it is possible that higher NAWM versus lesion tNAA is a reflection of *up-regulated* NAAG in NAWM as well as loss of NAAG (and NAA) in lesions and could be interpreted as a response to disease activity (Vrenken et al., 2005).

### 4.3 No differences in NAA or Inositol between lesions and NAWM in AQP4Ab-NMOSD patients

Within our four AQP4Ab-NMOSD patients, no differences were found between lesion and control sites for NAA, tNAA or Ins. Interpretation of these results is of course difficult given the small number of AQP4Ab-NMOSD patients in this study. However, to our knowledge there is only one other study describing AQP4Ab-NMOSD MRS findings in central nervous system lesions and that study focuses solely on the spinal cord (Ciccarelli et al., 2013). Ciccarelli and colleagues found lower Inositol in AQP4Ab-NMOSD lesions relative to MS lesions which in turn had lower concentrations than healthy controls, and they hypothesised that this demonstrated astrocyte loss. There are no brain studies of AQP4Ab-NMOSD lesions versus NAWM for appropriate comparison and assumptions about AQP4Ab-NMOSD brain lesion metabolites from spinal cord data should be made with caution, especially given that no AQP4Ab-NMOSD control site was sampled in Ciccarelli’s study and no healthy control group was sampled in ours. It must also be noted that both ours and Ciccarelli’s studies have low numbers of AQP4Ab-NMOSD participants (4 and 5, respectively).

### 4.4 tNAA higher in NAWM in MS than AQP4Ab-NMOSD patients

There were no differences for *a priori* tests comparing AQP4Ab-NMOSD and MS lesion and control sites. However a post-hoc non-parametric t-test performed after unexpected differences were noted in plotted data revealed tNAA to be greater in MS compared to AQP4Ab-NMOSD NAWM, contrary to our hypothesis. Low tNAA in MS in contrast to AQP4Ab-NMOSD NAWM is usually offered as support for the hypothesis that MS pathogenesis involves a chronic extra-lesional neurodegenerative processes that is absent from AQP4Ab-NMOSD pathology (Huda et al., 2019). However, this finding has been challenged by Vrenken et al who found that the only significant difference between healthy controls and MS NAWM was a difference in NAAG (and not tNAA or NAA), and that NAAG wasn’t *lower* in NAWM of MS patients but was instead *raised* compared to controls (Vrenken et al., 2005). This explanation better fits with our data and is supported by a recent study of healthy participants, in whom absolute tNAA NAWM concentrations are lower than in MS NAWM here, and more in line with our AQP4Ab-NMOSD tNAA NAWM values (∼8.7 mmol/L for 20-60 year olds) (Ding et al., 2016).

Contrary to our initial hypothesis, we did not find greater Inositol in MS lesions compared with AQP4Ab-NMOSD lesions. It is not clear why this might be, and interpreting a null result in an n of 4 should be treated with caution, but it may be that this reflects the known pathology of some chronic AQP4Ab-NMOSD lesions where a period of gliosis supervenes over the astrocytic death of the acute lesion stage (Lucchinetti et al., 2014). MRS Inositol has been previously suggested as a potential differentiator for MS and AQP4Ab-NMOSD because astrocytes are reduced pathologically in AQP4Ab-NMOSD and increased in MS (Geraldes et al., 2018), but this assumption rests on data from the only study besides ours to have performed MRS in AQP4Ab-NMOSD lesions, and that study was in spinal cord lesions not brain lesions (Ciccarelli et al., 2013).

One explanation for the different Inositol concentrations between Ciccarelli’s study and the results presented here is that we have studied lesions of different ages: our study examined older lesions more likely to be in the gliotic stage (4-24 months vs 43-116 months). It is also possible that spinal cord lesions are in general more destructive than brain lesions, leading to astrocyte loss, whereas brain lesions are commonly more subtle, non-demyelinating, non-necrosing and sometimes reversible (indeed, historically, brain lesions were thought to be atypical of AQP4Ab-NMOSD)(Lucchinetti et al., 2014; Pittock et al., 2006). Finally, AQP4Ab-NMOSD lesions in the cord are centred on grey matter, while those in the brain are found primarily white matter, which may be an important factor in gliosis (Lucchinetti et al., 2014).

### 4.5 Ins:NAA correlates with disability

We also found a correlation between Ins:NAA and disability (as assessed by EDSS), which was significant on post-hoc testing for both disease groups combined and for AQP4Ab-NMOSD alone (Supplemental Figure 2). This correlation suggests that higher Ins:NAA relates to worse clinical score. Consistent with this finding, NAWM Ins:NAA has been shown in one study of MS patients to predict subsequent clinical disability, with higher Ins:NAA predicting greater EDSS score increase (Llufriu et al., 2014). It has been hypothesised that the CNS immune process in MS leads to an increase of inositol and the neurodegeneration causing long-term disability results in reduced NAA, hence Ins:NAA is greater in more destructive and longer-lasting disease (Llufriu et al., 2014; Miller DH, 2014). We found a similar association of Ins:NAA and EDSS across the whole group, but this was primarily driven by the strong relationship between Ins:NAA and EDSS in AQP4Ab-NMOSD NAWM (albeit in n=4). This is surprising, as the received wisdom is that AQP4Ab-NMOSD NAWM is relatively free from damage, at least outside the optic nerve and cortico-spinal tracts (Aboul-Enein et al., 2010; Bichuetti et al., 2008; De Seze et al., 2010; Matthews et al., 2015). However, even if only small differences in NAWM NAA and Inositol occur, provided the differences decrease in magnitude with increasing EDSS, Ins:NAA will increase proportionally with EDSS. In our four AQP4Ab-NMOSD patients we found no-change or a slight increase in NAA with EDSS (i.e. no evidence of neuronal loss in NAWM in line with existing literature), along with a proportionally greater increase in Ins.

### 4.6 Limitations

Our study sought to compare AQP4Ab-NMOSD and MS neurochemicals using MRS, but has some limitations. Firstly, no healthy control population was used as a comparator for NAWM brain voxels. Instead, patients’ own NAWM was used as a control site to compare with lesion sites. As such, it is impossible to be sure whether the differences described above are driven by pathological increases or decrease or both, except through comparison with previously published values, which have invariably used different techniques and assessed different anatomical locations. It is thus possible that a structured difference in sampling location by disease type has biased the findings, given that we know MRS measures differ slightly across different lobes of the brain (Ding et al., 2016). Further, there is MRS and MRI diffusion imaging evidence of mirror changes (tNAA:tCr and apparent diffusion coefficient, respectively) that occur contralaterally to sites of lesions in multiple sclerosis (Stefano et al., 1999; Werring et al., 2000). These could conceivably reduce the magnitude of lesion versus NAWM differences investigated here, or hide them altogether. Secondly, lesions were all at least 3 months old (i.e. chronic) at the time of assessment. This makes our results comparable with most but not all previous MS MRS lesion studies but means little insight can be gained into changes with lesion progression. Finally, AQP4Ab-NMOSD disproportionately affects Afro-Caribbean individuals, whereas MS is commonly considered a disease of Caucasians. As such, matching ethnicity is difficult for studies comparing these two patient groups. It is not clear what effect, if any, this would have on the data.

## 5. Conclusion

Here we present results from an early, comprehensive metabolite profile of MS and AQP4Ab-NMOSD chronic lesions and normal appearing white matter acquired using magnetic resonance spectroscopy (MRS) at 7T. The study utilises an optimised methodology, including correction for multiple T2-water relaxation times, and our results are broadly in line with previous MRS studies in neurodegerative conditions, but serve to highlight some under-explored subtleties in MRS profiles, such as the absence of Inositol concentration differences in AQP4Ab-NMOSD brain lesions versus NAWM and the important influence of NAAG differences between lesions and normal appearing white matter. We hope that the technique described here will be highly relevant for future 7T MRS studies of this sort.

## Data Availability

Due the clinically sensitive nature of the data it is have not been made freely available. However, should you or your organisation have an interest in acquiring this data for the purpose of furthering the understanding of multiple sclerosis and neuromyelitis optica, please get in touch with the corresponding author.

## Acknowledgements

This work was funded by the MS Society (grant number 858/07) and the Guthy Jackson Charitable Foundation. The research was additionally supported by the National Institute for Health Research (NIHR) Oxford Biomedical Research Centre, and the NIHR Oxford Health Biomedical Research Centre. The Wellcome Centre for Integrative Neuroimaging is supported by core funding from the Wellcome Trust (203139/Z/16/Z). GT holds an Institutional Strategic Support Fund (ISSF) Fellowship funded by the Wellcome trust and received funding from the Economic and Social Research Council’s (ESRC’s) postdoctoral fellowship programme prior to the current post. CJS holds a Sir Henry Dale Fellowship, funded by the Wellcome Trust and the Royal Society (102584/Z/13/Z).

## Declaration of Interest

GT reported no declarations of interest.

YK reported no declarations of interest.

RM reported no declarations of interest.

SM reported receiving travel grants from Biogen, Novartis, Bayer, Merck & Co, Roche, and Almirall and honorarium from Biogen for advisory work.

AW reported no declarations of interest.

AC reported no declarations of interest.

RE reported no declarations of interest.

GDL is supported by the NIHR Biomedical Research Centre (BRC), Oxford and has research funding from the Oxford BRC, MRC(UK), UK MS Society, and National Health and Medical Research (Australia). GD has received travel expenses from Bayer Schering, Biogen Idec, Genzyme, Merck Serono, Novartis, American Academy of Neurology, and MS Academy, and honoraria as an invited speaker for Novartis, American Academy of Neurology, and MS Academy.

AW reported no declarations of interest.

MC reported no declarations of interest.

IT reported no declarations of interest.

JP reported no declarations of interest.

CS reported no declarations of interest.

UE reported no declarations of interest.

## Abbreviations

Asc: ascorbate
Cr: creatine
GABA: gamma aminobutyric acid
Glc+Tau: glucose and taurine
Gln: glutamine
Glu: glutamate
Glx: Gln + Glu
GPC: glycerophosphocholine
GSH: glutathione
Ins / Inositol: *myo*-inositol
NAA: N-acetylaspartate (i.e. *not* including NAAG)
NAAG: N-acetylaspartylglutamate
PCr: phosphocreatine
PE: phosphorylethanolamine
Tau: taurine
tCho: total choline (GPC + Cho)
tCr: total creatine (Cr + PCr)
tNAA: total N-acetylaspartate (NAA + NAAG)

---

**Supplemental Figure 1.**
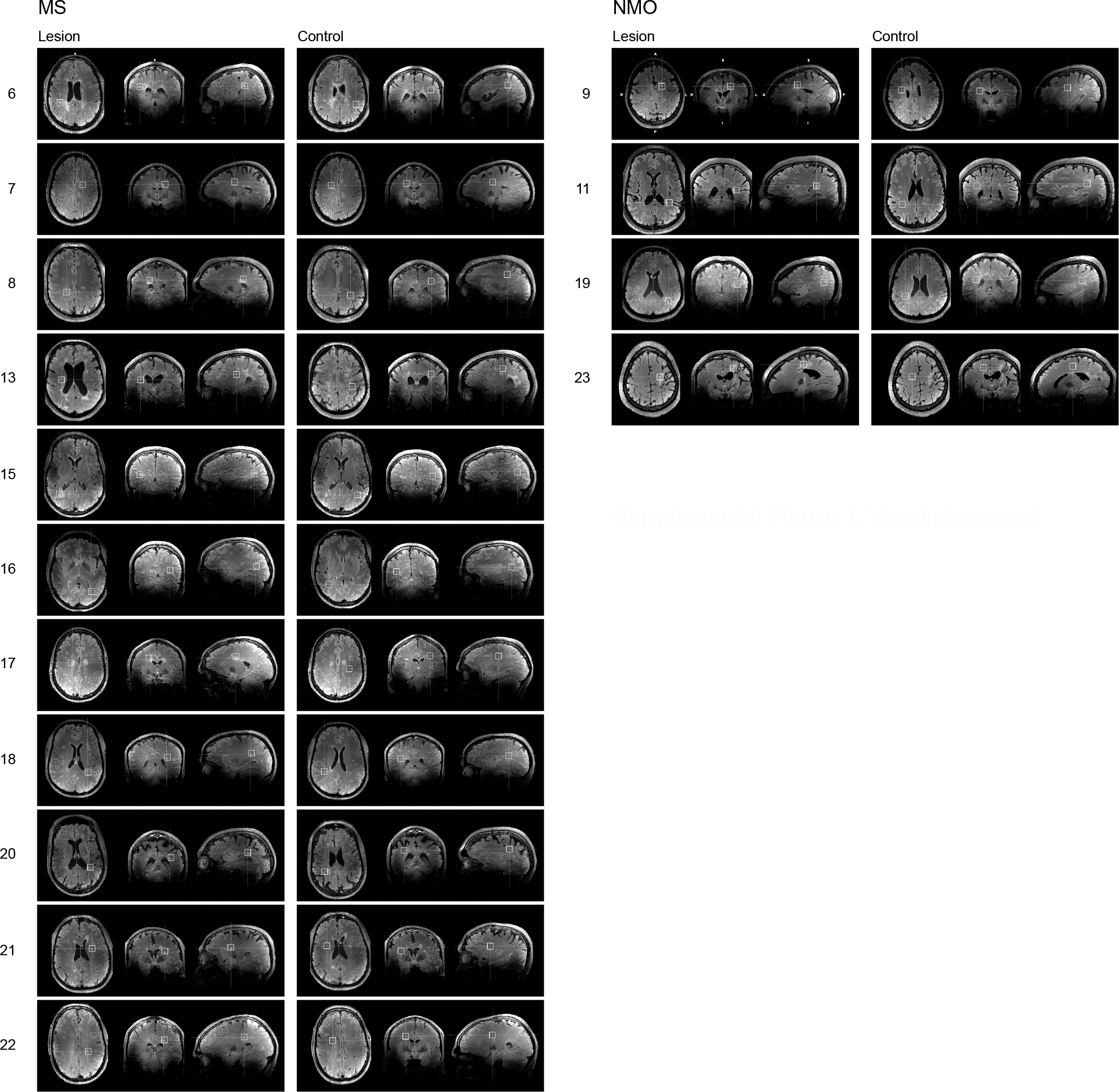
Voxel placement.

**Supplemental Figure 2.**
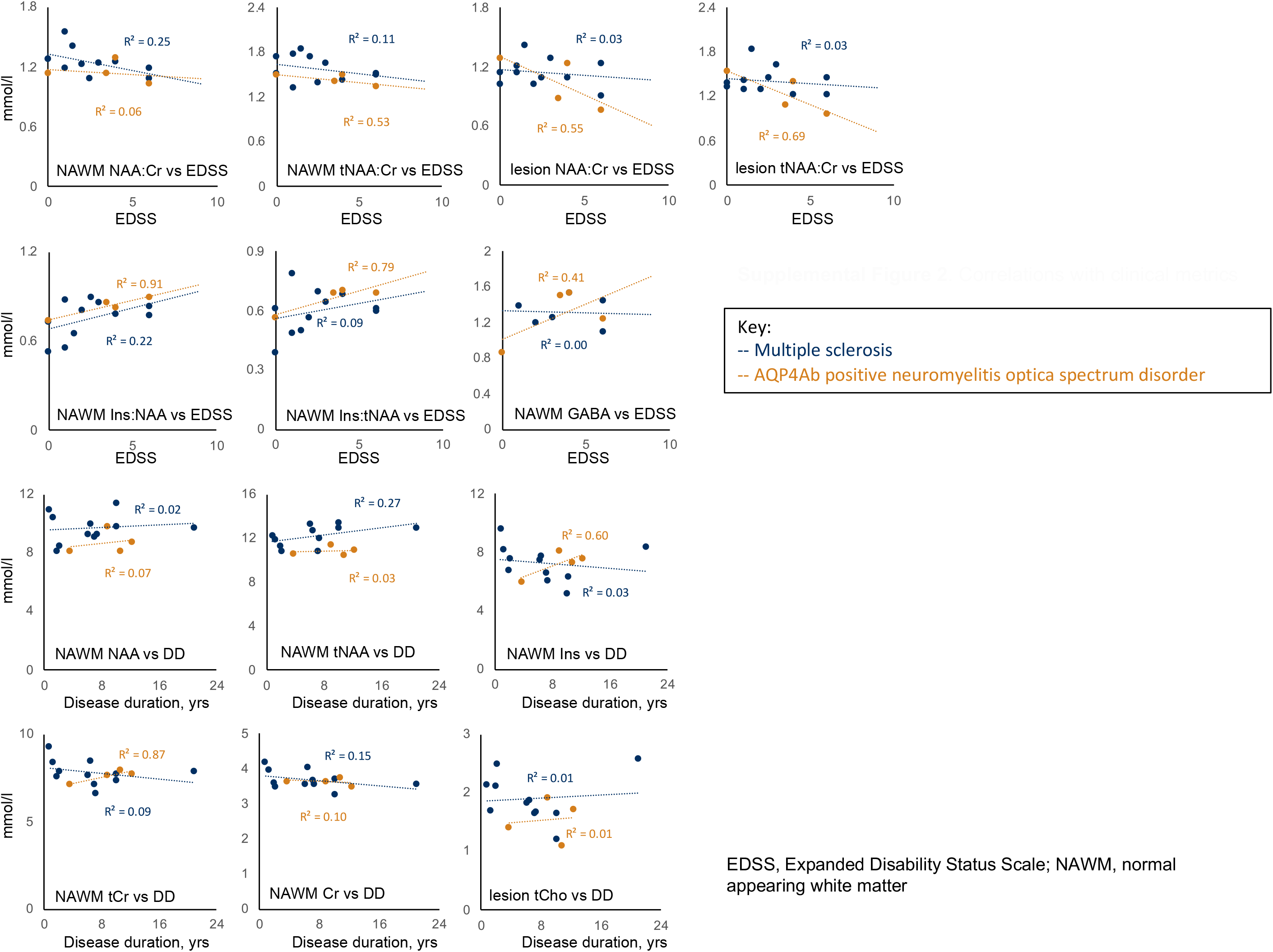
Correlations with clinical metrics.

